# RELATIONSHIP BETWEEN PARTICIPATION IN DAILY LIFE ACTIVITIES AND PHYSICAL ACTIVITY IN STROKE SURVIVORS: A PROTOCOL FOR A SYSTEMATIC REVIEW AND META-ANALYSES-BASED DATA SYNTHESIS

**DOI:** 10.1101/2023.05.05.23289233

**Authors:** Cristina de Diego-Alonso, Julia Blasco-Abadía, Almudena Buesa Estellez, Rafael Giner-Nicolás, María Pilar López-Royo, Patricia Roldán-Pérez, Víctor Doménech-García, Pablo Bellosta-López, Natalie A Fini

**Affiliations:** Universidad San Jorge. Campus Universitario, Autov. A23 km 299, 50830. Villanueva de Gállego, Zaragoza, Spain; Department of Physiotherapy, Melbourne School of Health Sciences, University of Melbourne, Melbourne, Victoria, Australia

**Keywords:** physical activity, participation, activities of daily living, stroke, rehabilitation, relationship, protocol, systematic review, meta-analysis

## Abstract

**Background:** Stroke survivors undertake low levels of physical activity and participation in daily life activities, but there are no reviews on the relationship between physical activity and participation. This systematic review and meta-analyses-based data synthesis will aim to cross-sectionally and longitudinally describe and estimate the relationship between participation in daily life activities and physical activity in stroke survivors.

**Methods and analysis:** This protocol follows the Preferred Reporting Items for Systematic Review and Meta-Analysis Protocols (PRISMA-P). Six databases (PudMed, Web of Science, Scopus, PEDro, SPORTDiscus, and Rehabilitation & Sport Medicine Source) will be searched using terms grouped into three categories: stroke, physical activity, and participation in daily life activities. Observational and experimental studies assessing participation and physical activity levels in adult stroke survivors in English or Spanish will be included. The methodological quality of the studies will be assessed independently by 2 investigators, with the Newcastle-Ottawa scale. Study and sample characteristics, and physical activity and participation outcome data will be extracted. Additionally, if available, repeated measures and correlation values between physical activity and participation outcomes will be extracted. The Hedges-Olkin method will be used for pooling correlation values and a random-effects meta-analysis following restricted maximum likelihood estimation for pooling effect sizes of those studies prospectively assessing the same measures of participation and physical activity. Subgroup analyses will be performed if a sufficient number of studies are found. Heterogeneity between studies results will be analyzed using I^2^ statistics, and publication bias will be examined using funnel plots and Egger’s tests.

**Discussion:** This will be the first systematic review with meta-analysis-based data synthesis to provide information on the relationship between physical activity and participation in stroke survivors. Findings are likely to inform, the design of health prevention protocols and the development of healthy behavior change interventions.

## INTRODUCTION

Stroke is the second leading cause of death in developed countries and, according to World Health Organization (WHO) projections, one in four people will suffer a stroke throughout their lifetime [1]. In survivors, stroke is the third leading cause of disability worldwide [2] as it often leads to serious sequelae including sensory-motor, cognitive and emotional impairments [3], which require neurorehabilitation [4]. For example, two-thirds show difficulty in walking independently, which is maintained beyond 3 months post-stroke [5]. These post-stroke sequelae generate limitations in activities, and restrict participation in daily life activities such as social or leisure activities [6, 7]. While inmmediate post-stroke sequelae might be inevitable, the subsequent sedentary lifestyle often observed in stroke survivors is believed to be modifiable [8]. Therefore, participation and physical activity (PA) are key targets for a healthy lifestyle in stroke survivors.

Participation is defined by the International Classification of Functioning, Disability and Health (ICF) as a person’s involvement in life situations, which means an interaction between the individual’s health condition and contextual factors (i.e., environmental and personal factors) [9, 10]. In addition, participation implies the performance of meaningful activities, habits, role performance, and social and community involvement [11]. Restoration of participation and fuctional levels is the main goal in occupational therapy [12]. However, the evidence regarding factors influencing the recovery of participation in daily life activities is limited. Commonly work, leisure, and domestic activities are the most affected after stroke [13, 14].

Physical activity is defined by the WHO as “any bodily movement produced by skeletal muscles that requires energy expenditure”, therefore, includes all movement during leisure time, for transport to get to and from places, or as part of a person’s work and domestic duties [15]. Physical activity can involve different intensities, such as light, moderate, and vigorous-intensity PA [15]. Following stroke, survivors are commonly inactive and have significantly increased sedentary lifestyles [16-18], spending long periods without activity, which may be related to the effects of the stroke [19]. In essence, post stroke PA levels are lower than that recommended by the WHO for preventing diseases such as stroke [20].

It has been observed that a greater capacity for functional independence implies greater participation, with the ability to walk and drive, as well as having an extensive social network, being considered enablers [21]. However, it has been shown that despite increased physical functioning, most people continue to have participation restrictions in social and leisure [21], work [22], and housekeeping activities [23], which generates a high level of concern for stroke survivors [24].

A previous review recommended that studies must include several complementary assessment tools to globally record the level of participation and PA to establish a complete correlation between these variables [25, 26]. To date, no systematic review has been conducted to synthesize the evidence on the relationship between participation and PA in stroke survivors. Therefore, the objective of this systematic review will be to cross-sectionally investigate and estimate the relationship between participation in the different activities of daily living and PA. Furthermore, a secondary aim will be to quantify participation and PA level trajectories and describe whether they follow a similar pattern over time.

## MATERIALS AND METHODS

### Study design and registration

This protocol follows the Preferred Reporting Items for Systematic Review and Meta-Analysis Protocols (PRISMA-P) [27] and it was registered in the International Prospective Register of Systematic Reviews (PROSPERO) database (CRD42022360711).

### Eligibility criteria

#### Study types

Observational (e.g., cross-sectional, case-control, cohort studies) and experimental studies (e.g., randomized control trials, randomised cross-over trials), evaluating participation and PA levels in stroke survivors will be included. Only articles with full-text is available and published in a peer-reviewed journal in English or Spanish languages will be included. The search will not be limited by date of publication, but only studies with a sample size higher than 10 participants will be included.

Studies with a prospective design including repeated measures of participation and PA measures across a follow-up period will be selected for inclusion in the data synthesis to quantify trajectories over time.

#### Participants characteristics

Studies recruiting adult participants (≥18 years) with stroke diagnosis regardless of time since stroke or severity of stroke sequelae, the aetiology, sex, or geographical location will be included. Studies with mixed populations with composite data will be excluded, unless stroke data is able to be extracted separately.

#### Characteristics of the outcome measures

Studies reporting PA data (e.g., minutes of PA at different intensities, caloric expenditure per PA, steps, sitting/standing time) measured by objective devices (e.g., podometers, accelerometrs), self-reported questionnaires or observational tools and, degree of participation in at least at least one area of daily living activity according to the occupational therapy framework: basic activities and instrumental activities of daily living, rest and sleep, education, work, play, leisure, or social participation [11] will be included. Studies only including physical performance tests will be excluded, as these cannot be considered as direct indicators of PA.

### Data sources and searches

A search will be carried in six electronic databases (PudMed, Web of Science, Scopus, PEDro, SPORTDiscus and Rehabilitation & Sport Medicine Source). Search terms will be grouped into three categories: stroke-related words, PA terms, and participation in daily life activities. The PubMed search strategy is provided in S1 Appendix.

To identify additional records, a detailed review of the bibliographic references included in the full-text reviewed articles will be performed.

### Study selection

After removing duplicates, the first step will consist of screening studies by titles and abstracts. Next, full texts will be reviewed and evaluated by applying the eligibility criteria. Studies will be selected at each step by 2 independent researchers (CDA and ABE), and, in case of disagreement, a consensus will be sought by a third researcher (PBL).

### Evaluation of the methodological quality of the studies

Two researchers (CDA and PRP) will independently examine the methodological quality of the studies using the Newcastle-Ottawa scale [28]. A third researcher (PBL) will verify the assessments and resolve discrepancies if any arise.

The Newcastle-Ottawa scale evaluates seven to eight items categorized into three criteria (selection, comparability, and exposure or outcome) with a maximum score of 9 (10 in the case of cross-sectional studies). Articles scoring at least 7 will be classified as “high quality”, 4-6 as “fair quality”, and less than 4 as “poor quality” [28]. Due to the observational nature of this review, clinical trials will be also evaluated with the Newcastle-Ottawa scale for prospective studies.

### Data collection

Two independent researchers (CDA and RGN) will extract data from the selected studies using a standardised data extraction sheet, while a third and fourth researcher (PBL and JBA) will create independent data bases, verify that the extracted data match, and resolve any discrepancies.

The following data will be collected: general study information (title, authors, year of publication); sample and subgroup characteristics (sample size, mean age and sex, time since stroke, stroke severity); study characteristics (research design and country); outcome measures (timing of measurement, PA objective device, PA self-reported assessments, participation assessments); intervention, if any; outcome data (PA and participation). In addition, main results, including correlation or association findings between participation and PA outcomes will be extracted when possible.

Furthermore, when possible, the mean and standard deviation at baseline (first assessment available) and follow-up endpoints from prospective studies will be extracted for participation and physical activity outcomes. In cases where the data is not reported directly in an article, up to three attempts will be made to contact the study authors via emailto obtain the data. Where available, medians and interquartile ranges will be extracted and transformed into mean and standard deviation [29].

### Data synthesis

When there is at least two studies that report a correlation between the same participation and PA outcomes, the weighted summary of correlation coefficients under the random effects model will be calculated using the Hedges-Olkin method, based on a Fisher Z transformation of the correlation coefficients [30].

When a minimum of two prospective studies with at least two independent cohorts examining the same participation and PA outcomes, repeated-measures, Hedges’ *g* effect sizes will be calculated (i.e., the result of subtracting the baseline mean minus the follow-up mean, divided by the averaged standard deviation weighted by sample size) [31] and pooled with a random-effects meta-analysis following a restricted maximum-likelihood estimation [32]. Absolutes values of effect size will be considered small (*g* ≥ 0.20 & <0.50), medium (*g* ≥ 0.50 & <0.80) or large (*g* ≥ 0.80) [33]. If sufficient studies are found, subgroup analysis will be performed according to areas of participation and/or PA measurement method (i.e., self-reported or objective measures). Moreover, age, sex, and/or methodological quality will be included in the analysis as moderators.

Heterogeneity between studies’ results will be investigated using I^2^ statistics with values >50% indicating substantial heterogeneity across studies [34]. Publication bias will be examined by using funnel plots and Egger’s tests [35].

## DISCUSSION

Recent estimates predict a high increase in the number of stroke survivors worldwide by 2040-2050 [2, 36-41]. This highlights the need to target participation in daily activities and PA levels for secondary prevention. A low level of PA is one of the main risk factors for relapse, which, together with the level of participation in daily life activities, is an indicator of the quality of life. Furthermore, a low PA level is related to the degree of independence andhas therefore an impact on health expenditure [42]. Considering the current situation and the demand from the WHO [43], it is urgent to update and synthesise the information available to date. Moreover, there is no systematic review with meta-analysis providing information on the relationship between PA levels and participation. To date these, data have been analysed separately[6, 44-48] in the stroke population.

This protocol describes the methodology that will be used to conduct the first systematic review with a meta-analysis-based data synthesis on the relationship between participation in daily life activities and PA in stroke survivors. The results obtained will potentially have a major impact on current knowledge by synthesising the information from all the publications to date, which will lay the foundations for future research and public health protocols in stroke survivors. Moreover, the results obtained will provide valuable insights into the important relationship between PA levels and participation which will help our understanding of this complex population and complex behaviours.

It should be noted that this systematic review will have some limitations. Research that uses a synonym for participation or PA may be missed by our searches. It is also possible that this review will not find a large number of high-quality publications that establish correlations between both elements.

## Data Availability

Deidentified research data will be made publicly available when the study is completed and published.

