## Supplementary Table 1 for "RELATIONSHIP BETWEEN PARTICIPATION IN DAILY LIFE ACTIVITIES AND PHYSICAL ACTIVITY IN STROKE SURVIVORS: A PROTOCOL FOR A SYSTEMATIC REVIEW AND META-ANALYSES-BASED DATA SYNTHESIS": Supplementary Material.pdf

**Appendix 1. SEARCH STRATEGY**

| **PUBMED** | **1# AND 2# AND 3#** |
|---|---|
| **1#**<br>**Stroke** | "Stroke"[Mesh] OR stroke[tiab] OR “cerebrovascular accident”[tiab] OR “cerebral vascular accident”[tiab] OR “cerebral vasospasm”[tiab] OR “cerebral bleed”[tiab] OR “post stroke”[tiab] OR poststroke[tiab] OR “cerebral ischaemia”[tiab] OR “brain ischemia”[tiab] OR “brain infarct”[tiab] OR “cerebral infarct”[tiab] OR “brain haemorrhage”[tiab] OR “cerebral haemorrhage”[tiab] OR “intracerebral haemorrhage”[tiab] OR “subarachnoid haemorrhage”[tiab] OR “intracranial haemorrhage”[tiab] OR “brain hemorrhage”[tiab] OR “cerebral hemorrhage”[tiab] OR “intracerebral hemorrhage”[tiab] OR “subarachnoid hemorrhage”[tiab] OR “intracranial hemorrhage”[tiab] |
| **2#**<br>**Physical Activity** | "Sedentary Behavior"[Mesh] OR “sedentary time”[tiab] OR “physical inactivity”[tiab] OR “physical activity”[tiab] OR walk* OR steps OR MVPA OR MPA OR “moderate to vigorous physical” OR LPA OR LIPA OR"light intensity physical activity" OR “metabolic equivalent of task” OR "Actigraphy"[Mesh] OR “activity monitor” OR “activity tracker” OR acceleromet* OR “pedometer” OR sensewear OR sense-wear OR activPAL OR “PAL 2” OR PAL2 OR IDEEA OR actigraph* OR actiwatch* OR stepwatch OR "actical" OR"activ8" OR"apple watch" OR"googlefit" OR"movemonitor" OR"step activity monitor" OR"StepWatch" OR"axivity" OR“motion logger*” OR motionlogger* OR fitbit ORIPAQ OR“International Physical Activity Questionnaire” ORPASE OR“Physical Activity Scale for the Elderly” ORPACE OR“Physician-based Assessment and Counselling for Exercise Score” |
| **3#**<br>**Participation in daily life activities** | “occupational area*” OR “daily life activities”[tiab] OR “daily life activities”[tiab] OR “everyday occupations”[tiab] OR ADL[tiab] OR “activities of daily living”[tiab] OR BADL[tiab] OR “basic activities of daily living”[tiab] OR IADL[tiab] OR “instrumental activities of daily living”[tiab] OR “self-care”[tiab] OR “leisure activities”[tiab] OR “social activities”[tiab] OR “community activities”[tiab] OR “community-dwelling”[tiab] OR “activities at home”[tiab] OR “social life”[tiab] OR sleep*[tiab] OR rest[tiab] OR “Occupational habit*” OR autonomy OR independence OR “occupational balance” OR “occupational satisfaction” OR “role performance” OR “community participation” OR “social participation” OR “return to work” OR “recovery activities” OR “meaningful activities” OR “participation restriction” OR “participation satisfaction” OR “role performance” OR “community reintegration” OR “community mobility” OR “behavioural mapping” OR ACS OR “activity card sort” OR SIS OR “stroke impact scale” OR BI OR “barthel index” OR FAI OR “frenchay activities index” OR “assessment of life habits” OR LHS OR “london handicap scale” OR FIM OR “functional independence measure” OR FAM OR “functional assessment measure” OR LIFE-H OR “assessment of life habits” OR RNLI OR “reintegration to normal living” OR ACS OR“Activity Card Sort” |
